# Using explainable machine learning to identify patients at risk of reattendance at discharge from emergency departments

**DOI:** 10.1101/2020.12.02.20239194

**Authors:** F. P. Chmiel, D. K. Burns, M. Azor, F. Borca, M. J. Boniface, Z. D. Zlatev, N. M. White, T. W. V. Daniels, M. Kiuber

## Abstract

Short-term reattendances to emergency departments are a key quality of care indicator. Identifying patients at increased risk of early reattendance could help reduce the number of missed critical illnesses and could reduce avoidable utilization of emergency departments by enabling targeted post-discharge intervention. In this manuscript we present a retrospective, single-centre study where we created and evaluated an extreme gradient boosted decision tree model trained to identify patients at risk of reattendance within 72 hours of discharge from an emergency department (University Hospitals Southampton Foundation Trust, UK). Our model was trained using 35,447 attendances by 28,945 patients and evaluated on a hold-out test set featuring 8,847 attendances by 7,237 patients. The set of attendances from a given patient appeared exclusively in either the training or the test set. Our model was trained using both visit level variables (e.g., vital signs, arrival mode, and chief complaint) and a set of variables available in a patients electronic patient record, such as age and any recorded medical conditions. On the hold-out test set, our highest performing model obtained an AUROC of 0.747 (95% CI : 0.722-0.773) and an average precision of 0.233 (95% CI : 0.194-0.277). These results demonstrate that machine-learning models can be used to classify patients, with moderate performance, into low and high-risk groups for reattendance. We explained our models predictions using SHAP values, a concept developed from coalitional game theory, capable of explaining predictions at an attendance level. We demonstrated how clustering techniques can be used to investigate the different sub-groups of explanations present in our patient cohort.

## Introduction

The use of emergency departments (EDs) has been growing steadily over the last decade^1,2^, which in turn has contributed to increased overcrowding and extended waiting times. These factors have been linked to increased rates of adverse outcomes^3,4^, and it is therefore important to investigate the most efficient ways of using the available resources and minimise their unnecessary use. One way this can be achieved is by minimizing short-term reattendances, which describe a situation where a patient presents to an emergency department within 72 hours of having been discharged. The number of short-term reattendances can be minimised by both delivering the highest levels of patient care, thereby reducing the chance of missed critical illness and injury at the initial attendance, and by mitigating reattendances for reasons at least partially unrelated to the initial ED attendance.

Research has shown there are several factors indicative of short-term reattendance risk including social factors (e.g., living alone)^5^, depression^6^, initial diagnosis^7^, and historical emergency department usage^8^. Knowledge of these risk factors is important to clinical staff when planning discharge, but this is unlikely the most optimal way of determining those at risk of suffering from a significant illness following erroneous discharge or patients in need of additional support in the community following discharge. Predictive models, available as a decision support tool at the point of discharge, able to identify patients at increased risk of short-term reattendance may be able to significantly reduce the number of reattendances. Predictions, and any associated explanations, could be used by clinical staff to help inform intervention (e.g., further diagnostic tests) or enrich discussions about a patients discharge plan.

Machine learning models are a class of predictive models which are well positioned to add value to emergency department processes. They are particularily powerful because they can ingest a large number of variables and learn complex interactions and statisical trends between these variables and associated outcomes, ultimately making highly accurate predictions of patient outcomes. The potential in using machine learning models to inform clinical care and provide high levels of personaliation, is evident from the large amount of research into the applications of machine learning in healthcare^9^. Explainable machine learning, in particular, is attracting attention because of the strong assurance needs of healthcare^10,11^ In the context of this work, research has shown that machine-learning models can use a large number of clinical and administrative variables to provide estimates of a patients short-term reattendance risk^12,13^. Explanation is also important in this context, as explanations can help to inform the patients care trajectory and guide post-discharge intervention plans. In this manuscript we discuss a machine-learning model, utilizing historical (coded, inpatient) discharge summaries, alongside contemporary clinical data recorded during emergency department attendances to identify patients at increased risk of short-term reattendance following an ED attendance. We explain our predictions by calculating SHAP values for our model output and cluster these explanations to explore the different sub-groups at risk of reattendance in our cohort.

## Methods

### Dataset curation

Our dataset featured an pseudonymized version of all attendances by adults to Southamptons Emergency Department (University Hospitals Southampton Foundation Trust) occurring between the 1st April 2019 and the 30th of April 2020. The data set included attendance level information such as the results of any near-patient observations and high-level information included in the standard UK Emergency Care Data Set (e.g., outcome, arrival mode, duration of visit, and chief complaint). In addition to attendance-level variables, our dataset includes variables extracted from a patients electronic health record maintained by the University Hospitals Southampton Foundation Trust. These variables included any recorded medical complaints, which were obtained by extracting ICD10 coded conditions included in historical discharge summaries. An example of the most frequently observed medical conditions are presented in Table 1, alongside a breakdown of patient ages in our cohort. In total our dataset included 14 variables: patient age (estimated from year of birth), number of emergency department attendances in the 30 days prior to the attendance, the chief complaint of the attendance recorded at registration and triage (e.g., abdominal pain), the patients mode of arrival, any recorded medical conditions, the count of the number of medical conditions a patient has at the time of their attendance, measured vital signs (temperature, pulse and respiration rate, systolic blood pressure, and blood oxygen saturation levels), the Manchester Triage System score, triage pain score, the primary triage discriminator, (coded) discharge diagnosis, and the hour of day and day of the week the attendance occurred. A full data schema is presented in Supplementary Table 1 and quantitative descriptors of the variables are presented in Supplementary Table 2.

**Table 1.** Breakdown of patient age and most frequently occurring conditions in the training and test sets. Conditions are extracted from their ICD10 codes. A given condition is only associated with a small fraction of attendances, but in total 40.0 % of attendances resulting in discharge have at least one associated condition. Patient age and conditions are extracted from their last attendance in the respective data set.

|  | Number of patients |  |
| --- | --- | --- |
|  | Train set (N=28,945) | Test set (N=7,237) |
| **Age groups** |  |  |
| 18-24 | 5,039 | 1,244 |
| 25-34 | 5,867 | 1,500 |
| 35-44 | 4,310 | 1,108 |
| 45-54 | 3,957 | 1,024 |
| 55-64 | 3,514 | 822 |
| 65-74 | 2,876 | 701 |
| 75+ | 3,382 | 838 |
| **Condition** |  |  |
| Hypertension | 2,911 | 681 |
| History of smoking | 1,805 | 442 |
| Asthma | 1,633 | 392 |
| Depression | 1,544 | 393 |
| Current Smoker | 1,436 | 359 |
| Type 2 diabetes | 1,081 | 278 |
| Hypercholesterolaemia | 895 | 201 |
| Osteoarthritis | 775 | 189 |
| COPD | 706 | 157 |

Our study cohort featured 54,015 attendances which resulted in discharge directly from the ED. Of these attendances we discarded those that resulted in planned reattendance (N=328) and those occurring after 01/02/2020, removing attendances which occurred during the COVID-19 pandemic. Remaining attendances (N=44,294) were randomly split at the patient level to create a hold-out test set (N=8,847) containing attendances from 20% of patients and the remaining attendances (N=35,447) were used as the training set (Figure 1).

**Figure 1.**
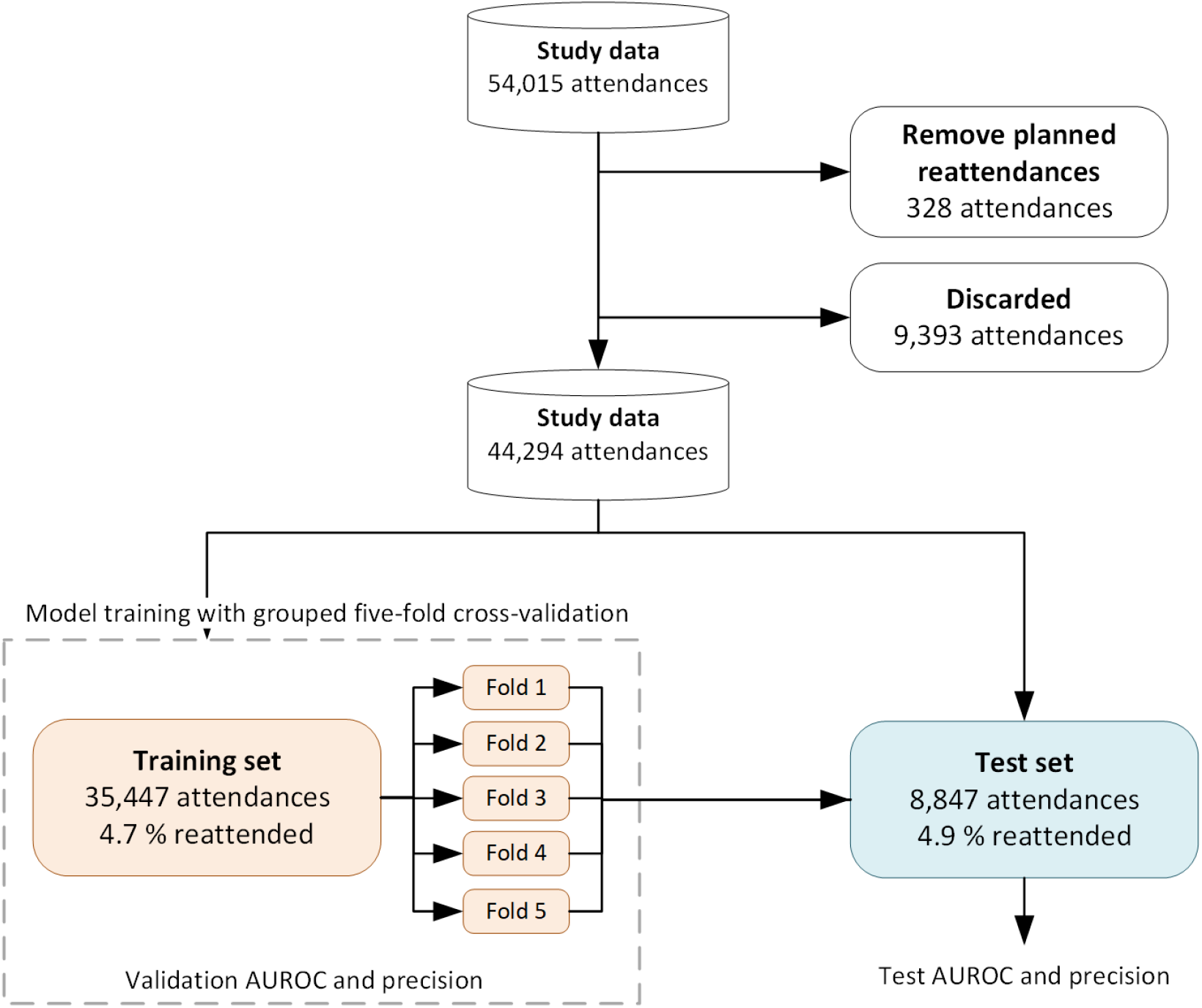
Segregation of the study data into the training and hold-out test set. Attendances which resulted in planned reattendances (328 attendances) were removed. All attendances (9,393 attendances) occurring after 01/02/2020 were also discarded to remove attendances which coincided with the COVID-19 pandemic. Reattendance rates (bottom row of rectangles representing training and test sets) display the observed 72-hour reattendance rate for each cohort. Models were trained using grouped (patients occur exclusively in either the training or validation folds) five-fold cross validation of the training set. Model hyperparameters were selected such that the validation AUROC (mean out-of-fold AUROC across the five folds) was maximized. The final model was the average output of the five models trained in the cross-validation and was evaluated on the hold-out test set.

### Ethics and data governance

Data was pseudonymized (and where appropriate linked) before being passed to the research team. The research team did not have access to the pseudonymisation key. Since the data was de-identified, Confidentiality Advisory Group approval was not needed and the study was approved by the NHS Health Research Authority (20/HRA/1102) without the need for informed consent because of the de-identified nature of the data. This study was approved by the University of Southamptons Ethics and Research governance committee (ERGO/FEPS/53164). Research was performed in the manner described in the protocol (v1) approved by the NHS Health Research Authority and conducted in accordance with relevant guidelines.

### Reattendance identification

Reattendances were identified by using the patient pseudo identifier to calculate the time to a patient’s next ED attendance. All reattendances (with the exception of planned reattendances, Figure 1) were considered, even if the reattendance was for a different reason to the original attendance. We then dichotomized the time to next attendance (return within 72 hours from the point of discharge), annotating each attendance with a binary indicator denoting whether it was followed by another attendance within 72 hours. This formulation allowed us to frame the predictive task as a binary classification problem.

### Predictive modelling

We used an extreme gradient boosted decision tree, as implemented in the XGBoost framework^19^, as our machine-learning model. Models were trained using five-fold cross validation of the training set, grouped at the patient level (Figure 1). Final predictions were the mean prediction of the fives models trained during the cross-validation process. Model hyperparameters were optimized by selecting the hyperparameters which maximized the validation AUROC (the mean performance of model on the five sets of out-of-fold samples). Hyperparameter search was performed using Bayesian optimization utilizing the Tree Parzen Estimator algorithm as implemented in the hyperopt Python library^17,18^. Medical conditions associated with a patient were included as a one-hot-encoded feature vector, the day of the week encoded using ordinal encoding, and all other categorical variables were encoded using target encoding^16^. The full encoding scheme is outlined in Supplementary Table 1.

To investigate the predictive ability of individual variables, under an independent variable assumption, we trained models using each distinct variable and evaluated them using five-fold cross validation on the training set. Models were trained (including hyperparameter tuning) using the previously described process. Next, we trained and evaluated a model using the three most predictive variables identified in this process and a model using all variables available at the point of discharge. During this analysis the standard error (95 % confidence) of the models performance across the five-folds were calculated. The final model (including all variables available at discharge) was then evaluated the hold-out test set, with 95 % confidence intervals calculated using bootstrapping (n=1000) of the hold-out test set. Models performance was evaluated using the Area Under the Receiving Operating Curve (AUROC) and the average precision under the precision-recall curve. Hyperparameters of our final model are presented in Supplementary Table 3.

We also evaluated the final model against patients *readmission* status (readmissions are a subset of reattendances for which the outcome was admission to hospital). This was performed without re-training the model and by evaluating the classifier with the ground truth outcome set to a patients readmission status.

### Model explainability

To explain the predictions of our final model we made use of the TreeExplainer algorithm implemented in the SHAP Python library^20–22^. TreeExplainer is an algorithm which calculates SHAP values (i.e, Shapley values), in an optimized manner for decision trees. SHAP values are a concept from coalitional game theory which treats predictive variables as players in a game and distributes their contribution to the predicted probability. SHAP values are particularly powerful as they meet the four desirable theoretical conditions of an explanation algorithm and can provide instance (i.e., attendance) level explanations^22^. Practically, for each attendance we have a scalar value for each variable used by the model which quantifies the contribution that variable had on the predicted reattendance risk. SHAP values of larger magnitudes indicate that a variable is of increased importance in determining the predicted reattendance risk. SHAP value can be negative (adding the variable reduces predicted reattendance risk) or positive (adding the variable increases the predicted reattendance risk).

To investigate the different explanations across the hold-out test set, we projected the SHAP values for all attendances in the hold-out test set into a two-dimensional (explanation) space using Uniform Manifold Approximation and Projection (UMAP)^23^. UMAP is a dimensionality reduction technique regularly used to visualise high-dimensional spaces in a low-dimensional embedding, such that global and local structure of the space can be explored^24,25^. Attendances which are closer in proximity in this space share a more similar explanation for their predicted reattendance risk. To investigate the characteristics of different sub-groups in the explanation space, we assigned each attendance to a cluster in this space using the DBScan algorithm^26^. Assignment was chosen by visual inspection and finer grained cluster assignment can be achieved by tuning the hyperparameters of the DBScan algorithm.

## Results

The results of our variable importance investigation are displayed in Table 2. Six variables (age, Manchester Triage System score, hour of day, vital signs, pain score, and arrival mode) were found to be weakly predictive of a patients 72-hour reattendance risk in isolation (AUROCs between 0.5 and 0.6, Table 2 models b-g). All other variables (Table 2 models h-n) were found to be moderately predictive (AUROC between 0.6 and 0.7) of 72-hour reattendance risk in isolation, with the exception of the day of the week the attendance occurred which was not predictive of a patients reattendance risk (model a, Table 2).

**Table 2.**
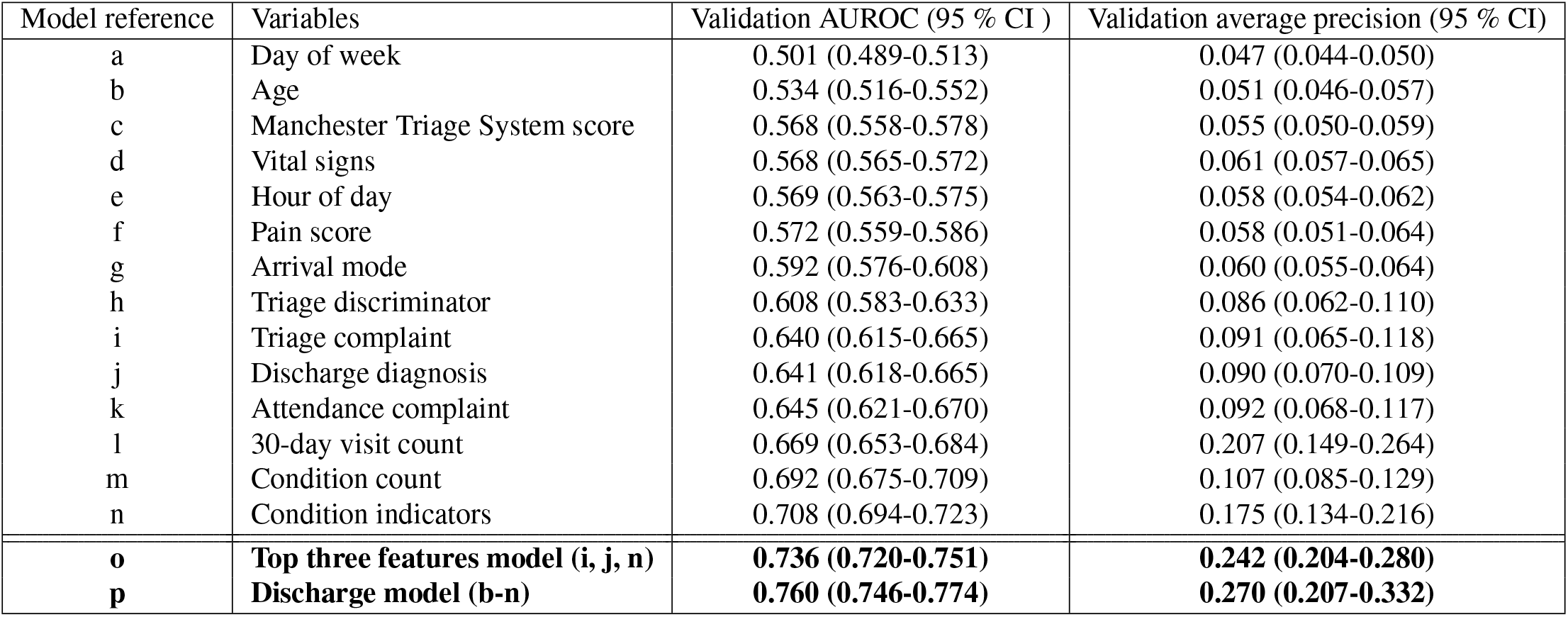
Performance on the validation set for models using individual variables (models a-n) and sets of variables (models o and p). Metrics are evaluated on the training set using grouped 5-fold CV at the patient level and we report the mean of the metric across the five validation folds. All models hyperparameters were tuned as described in the methods section to optimize the CV AUROC.

| Model reference | Variables | Validation AUROC (95 % CI ) | Validation average precision (95 % CI) |
| --- | --- | --- | --- |
| a | Day of week | 0.501 (0.489-0.513) | 0.047 (0.044-0.050) |
| b | Age | 0.534 (0.516-0.552) | 0.051 (0.046-0.057) |
| c | Manchester Triage System score | 0.568 (0.558-0.578) | 0.055 (0.050-0.059) |
| d | Vital signs | 0.568 (0.565-0.572) | 0.061 (0.057-0.065) |
| e | Hour of day | 0.569 (0.563-0.575) | 0.058 (0.054-0.062) |
| f | Pain score | 0.572 (0.559-0.586) | 0.058 (0.051-0.064) |
| g | Arrival mode | 0.592 (0.576-0.608) | 0.060 (0.055-0.064) |
| h | Triage discriminator | 0.608 (0.583-0.633) | 0.086 (0.062-0.110) |
| i | Triage complaint | 0.640 (0.615-0.665) | 0.091 (0.065-0.118) |
| j | Discharge diagnosis | 0.641 (0.618-0.665) | 0.090 (0.070-0.109) |
| k | Attendance complaint | 0.645 (0.621-0.670) | 0.092 (0.068-0.117) |
| l | 30-day visit count | 0.669 (0.653-0.684) | 0.207 (0.149-0.264) |
| m | Condition count | 0.692 (0.675-0.709) | 0.107 (0.085-0.129) |
| n | Condition indicators | 0.708 (0.694-0.723) | 0.175 (0.134-0.216) |
| **o** | **Top three features model (i, j, n)** | **0.736 (0.720-0.751)** | **0.242 (0.204-0.280)** |
| **p** | **Discharge model (b-n)** | **0.760 (0.746-0.774)** | **0.270 (0.207-0.332)** |

Medical condition history was included in two representations. The count of the number of historical conditions (model m, Table 2) obtained a validation AUROC of 0.692 (95 % CI: 0.675-0.709), reflecting that patients with a recorded medical history are more likely to reattend (8.3 % (95 % CI: 7.8-8.7 %) reattendance rate) than those who do not (2.3% (95 % CI: 2.1-2.5%) reattendance rate). When we included the full one-hot encoded matrix denoting whether the patient had a history of a specific condition, our model (model n, Table 2) obtained a validation AUROC of 0.708 (95 % CI: 0.694-0.723). The higher validation AUROC of the latter model suggests that different (medical) conditions are associated with differing degrees of reattendance risk.

The model that used the number of times a patient attended the emergency department in the 30-day prior to their current attendance (Table 2, model l) exhibited a validation AUROC of 0.669 (95 % CI: 0.653-0.684), agreeing with other studies that a patients previous emergency department usage is an important consideration when considering their reattendance risk^8^. Three models (models i, j, and k, Table 2) make use of coded information describing the primary reason for the emergency department attendance, recorded at three distinct points in time and by potentially different members of clinical and nonclinical staff. Making use of the chief complaint collected at either the point of registration or Triage, respective validation AUROCs of 0.645 (95 % CI: 0.621-0.670) and 0.640 (95 % CI: 0.615-0.665) could be achieved. At the point of discharge, the recorded diagnosis obtained a validation AUROC of 0.641 (95 % CI: 0.618-0.665). This demonstrates that different diagnoses are associated with differing degrees of reattendance risk and indicates that a high-level, coded description of the patients chief complaint is moderately predictive of reattendance risk, regardless of when it is recorded during the attendance.

Finally, models o and p in Table 2 present validation metrics for models trained using multiple variables. The model (model o in Table 2) using the three most predictive variables identified in our univariate importance study (Table 2) used the condition indicators, the condition count, and the number of times the patient visited the ED in the previous 30 days. We observed a validation AUROC of 0.736 (95 % CI: 0.720-0.751), demonstrating that models using multiple variables are more predictive of reattendance than a single variable. The model trained using all variables available at the point of discharge (model p in Table 2) increased the validation AUROC to 0.761 (95 % CI: 0.746-0.774).

The evaluation of our final model (model p) on the hold-out test set is presented in Figure 2. This model obtained an AUROC and average precision of 0.747 (95% CI : 0.722-0.773) and 0.233 (95% CI : 0.194-0.277) respectively on the hold-out test set. This indicate that while the model retained its moderate performance, there was a reduction in model performance between the validation and the hold-out test set. This is likely the result of the model overfitting the validation data (e.g., via the selection of hyperparameters to maximize model performance on the validation set), and is expected. In panel c of Figure 2 we display a confusion matrix of our model at a single configuration, where the threshold for dichotomization of the continuous predictions output by our model was chosen such that a recall of 0.15 was obtained. This graphic presents the performance of our model on the hold-out test set when used as a binary decision support tool with a low-recall configuration. Figure 3 summarizes the SHAP values of all attendances in the hold-out test set. In Figure 3 a the SHAP values for the ten most important variables (as determined by mean absolute SHAP values) are shown for each attendance. Visual inspection of this graphic provides insight into the trends our model has learned: the model associates anyone with a recorded medical condition as being at increased risk of reattendance and learned that some medical conditions represent a greater reattendance risk than others. For example, the model generally associates living alone with a higher reattendance risk than having a history of depression (the mean SHAP value is greater for those who live alone). In panels b and c of Figure 3 we plot the SHAP values across all attendances for two variables, the hour of day the attendance occurred and 30 day visit count. Visual inspection of panel b shows that the model associated the patients risk of reattendance with a periodic dependence on the hour of day the attendance occurred. By inspection of Figure 3 c, we can see that the model learned an approximately linear dependence between a patients reattendance risk and the number of times they have attended the emergency department in the last 30 days. It is important to note that these insights do not necessarily reflect actual risk factors for reattendance (since the model is an imperfect classifier) but only reflect the trends the model has learned to make its decisions.

**Figure 2.**
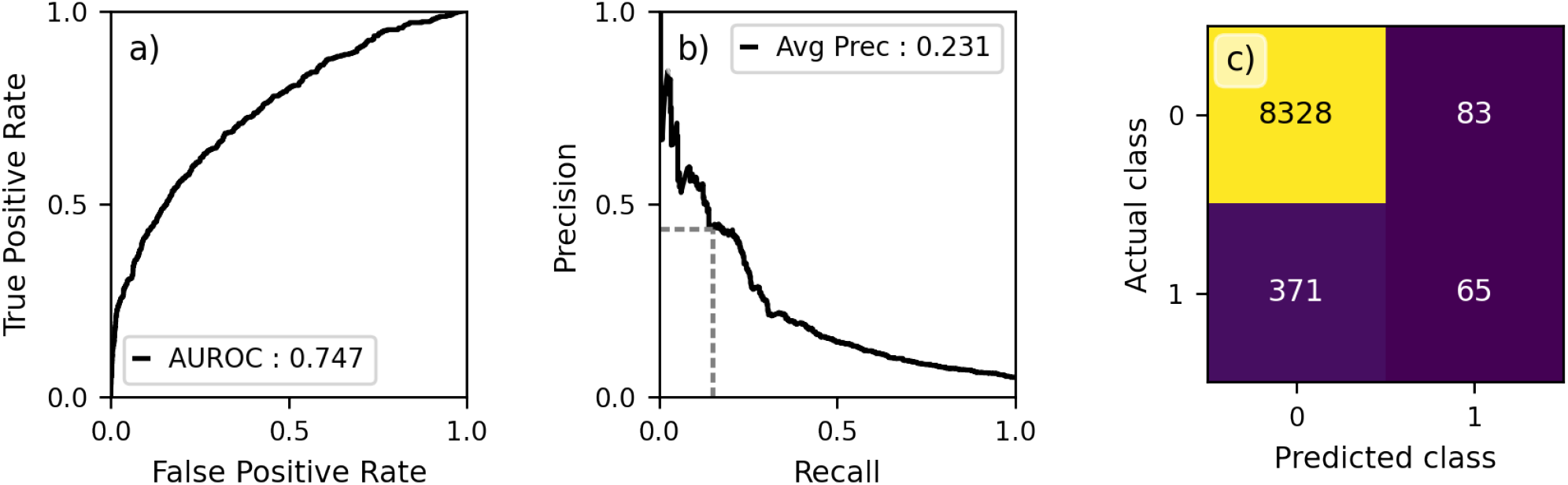
Performance of our model (model p in Table 2) evaluated on the hold-out test set. a) Receiver operating characteristic curve for models predictions. b) Precision recall curve, the dashed grey line shows the configuration evaluated in the confusion matrix in panel c. c) Confusion matrix for predictions dichotomized using a threshold chosen such that the recall is equal to 0.15 (dashed grey line in panel b). A class of 1 indicates the patient reattended the emergency department within 72 hours of discharge. Diagonal elements represent correct classifications and off-diagonal elements either False positives or negatives.

**Figure 3.**
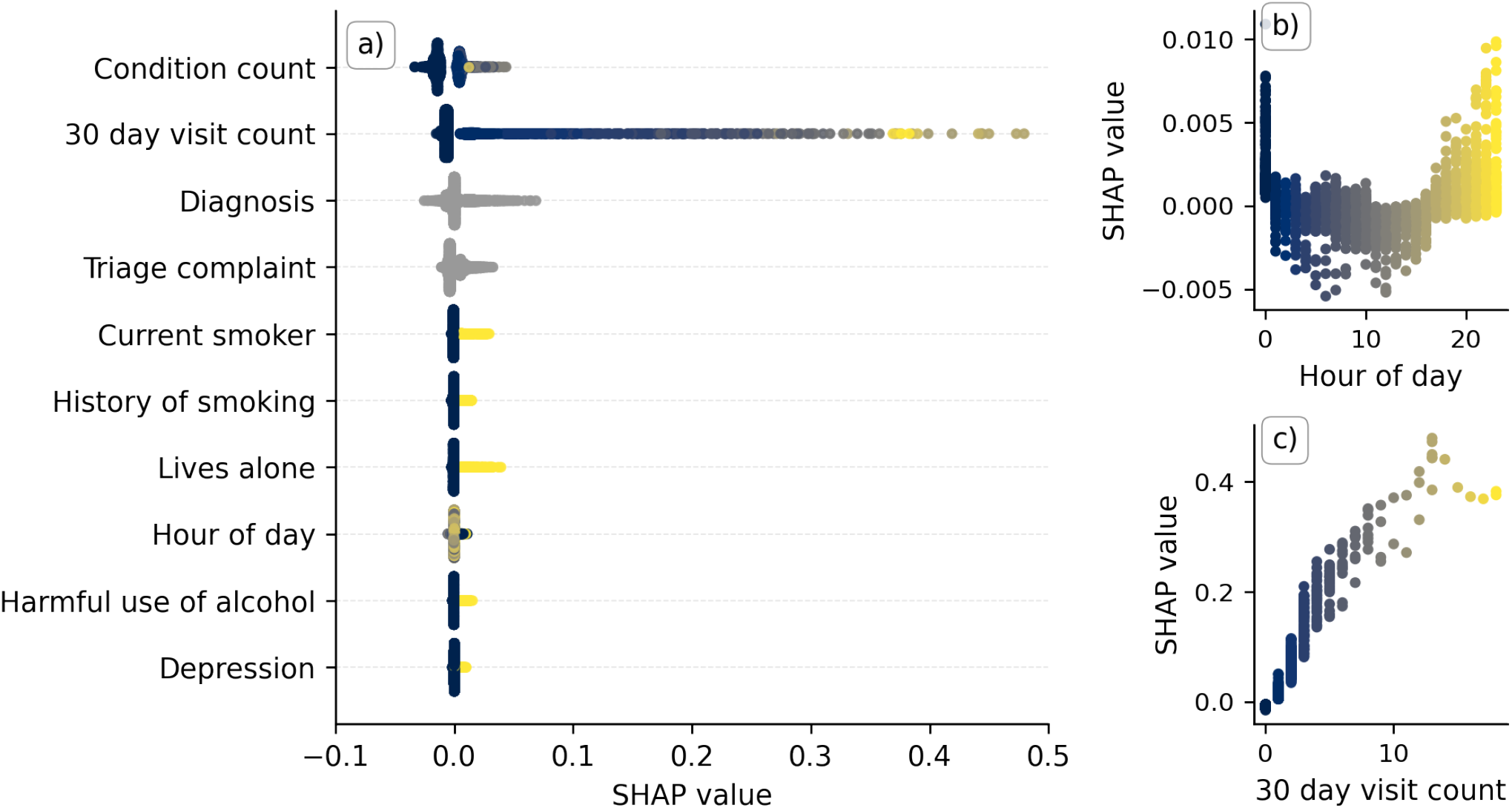
SHAP values for each attendance in the hold-out test set. a) Plot summarizing the SHAP values for the ten most important variables (by mean absolute SHAP value) for each attendance in the test set. They are ordered by the global impact the feature has on the explanation (equal to the mean absolute SHAP value of the feature across all attendances). For the binary variables (i.e., the condition indicators) this favours variables with a high number of occurrences (i.e., more common conditions), not necessarily those associated with the high reattendance risk. b) SHAP value against recorded hour of day of attendance registration (dots). c) SHAP value against number of emergency department visits in the 30 days prior to the given attendance (dots). In panels b and c vertical dispersion is the result of interaction with other variables. All panels are coloured by the magnitude of the respective variable for the given data point, with lighter colours indicating higher values (e.g., inspect panels b and c). Grey data points correspond to non-binary, nominal categorical variables and therefore have no natural ordering which could be used to colour the data points.

The projection of all SHAP values for attendances in the hold-out test set into the two dimensional explanation space (see Methods) is displayed in Figure 4. In panel a of Figure 4 we colour attendances by the reattendance risk predicted by our machine-learning model. The reattendance risk (colour) is relatively uniform, reflecting that the majority of attendances do not result in reattendance (the observed reattendance rate across the test set is 4.9 %). However, there are clear subgroups of the population identified to be at heightened risk of reattendance. In Figure 4 b we colour the attendances by the high-level cluster assignment obtained using the DBScan clustering algorithm. The characteristics of attendances in each clusters is displayed in Table 3. Visual inspection of Table 3 provides high-level insight into the logic of the machine-learning model. For example, for attendances assigned to cluster seven, on average, the most important variable for determining a patients reattendance risk is their 30-day visit count.

**Figure 4.**
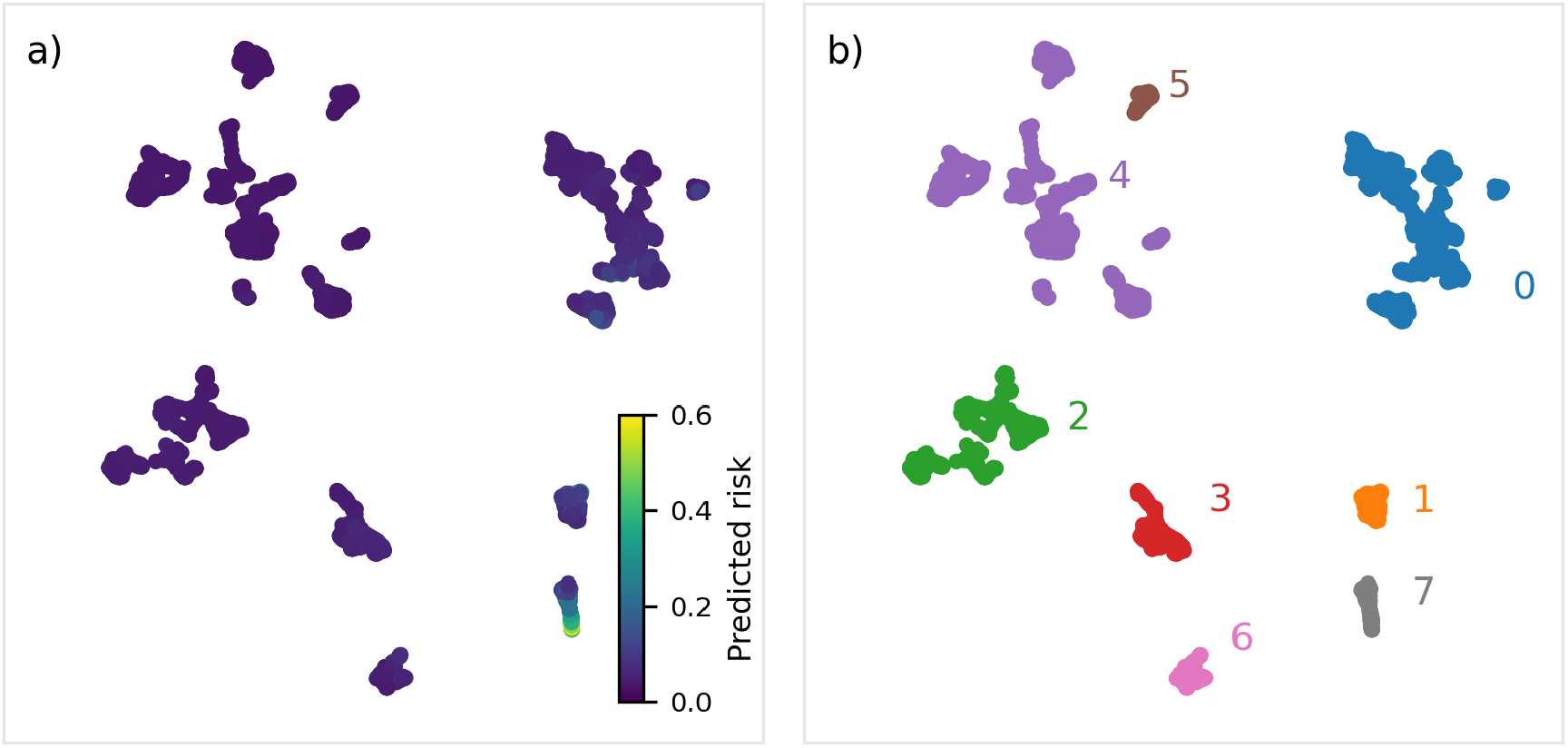
Embedding of the hold-out test set into a two-dimensional explanation space using the UMAP algorithm. a) All attendances in the hold-out test set visualised in the explanation space, colour indicates the predicted reattendance risk for the respective attendance. b) Attendances in the explanation space coloured by cluster assignment. Cluster assignment was performed with the DBScan algorithm^26^. Assignment was chosen by visual inspection and finer grained cluster assignment can be achieved by tuning the hyperparameters of the DBScan algorithm. The explanation embedding was created by clustering the SHAP values (explanations) for each attendance using the UMAP algorithm. Closer data points share a more similar explanation for their predicted reattendance risk. Descriptive properties of each cluster are displayed in Table 3.

**Table 3.** Properties of the attendances assigned to each of the explanation clusters (Figure 4). The count column displays the number of attendances in a given cluster. The age, reattendance rate, predicted reattendance rate, condition count, and 30-day visit count column display the mean of the respective variable for all attendances in the cluster. The final three columns display the most important variables in making a decision, averaged across all attendances in a given cluster.

| Cluster | Count | Age (years) | Reattendance rate (95 % CI) | Predicted reattendance rate (95 % CI) | Condition count | 30-day visit count | Most important variables (mean absolute SHAP values) |  |  |
| --- | --- | --- | --- | --- | --- | --- | --- | --- | --- |
|  |  |  |  |  |  |  | 1<sup>st</sup> | 2<sup>nd</sup> | 3<sup>rd</sup> |
| 0 | 2,723 | 55.4 | 5.9 (5.1-6.9) | 6.8 (5.0-8.7) | 2.9 | 0.0 | 30-day visit count | Triage complaint | Condition count |
| 1 | 446 | 52.81 | 8.5 (6.3-11.5) | 10.6 (7.1-14.0) | 3.5 | 1.0 | 30-day visit count | Medical history | Triage complaint |
| 2 | 1,363 | 42.45 | 2.6 (1.9-3.6) | 4.6 (4.4-5.0) | 0.0 | 0.0 | Condition count | 30-day visit count | Triage complaint |
| 3 | 671 | 40.9 | 4.0 (2.8-5.79) | 5.6 (5.0-6.3) | 0.0 | 0.0 | Condition count | Diagnosis | 30-day visit count |
| 4 | 2761 | 41.5 | 1.8 (1.3-2.3) | 3.8 (3.6-4.0) | 0.0 | 0.0 | Condition count | 30 day visit count | Triage complaint |
| 5 | 221 | 40.5 | 0.0 (0.0-1.7) | 3.6 (3.4-3.7) | 0.0 | 0.0 | Condition count | 30-day visit count | Triage complaint |
| 6 | 321 | 39.7 | 5.6 (3.6-0.9) | 5.9 (4.7-7.0) | 0.0 | 1.0 | Condition count | 30-day visit count | Triage complaint |
| 7 | 341 | 42.3 | 31.1 (26.4-36.2) | 27.2 (13.6-40.7) | 3.3 | 3.75 | 30-day visit count | Medical history | Condition count |

When evaluated on the readmission status (whether a patient reattended within 72 hours *and* was subsequently admitted to hospital) of attendances in the hold-out test set our model achieved an AUROC of 0.804 (95 % CI: 0.774-0.832) and an average precision of 0.044 (95 % CI: 0.030-0.063). Reflecting that the model demonstrated moderate discriminative ability in identifying the subset of reattendances which resulted in admission.

## Discussion

Our final 72-hour reattendance risk model achieved an AUROC of 0.747 (95% CI : 0.722-0.773) and an average precision of 0.233 (95% CI : 0.194-0.277) on the hold-out test set. Qualitatively, our model can use variables describing an emergency department attendance and a view of a patients medical history to predict their reattendance risk with moderate performance.

We calculated SHAP values to provide an explanation of our predictions at an attendance level (Figure 3 and Supplementary Figure 2) and projected these explanations into a two-dimensional space (Figure 4). We used this low-dimensional embeddings to identify different patient sub-groups at risk of reattendance.

Our final model (model p in Table 2) makes use of all variables in our dataset available at the point of discharge. This set of variables is not necessarily the optimal set of variables for a reattendance predictor, it is plausible the variable set contains obselete information because of correlations between variables. High correlation between variables is expected for clinical data. For example, it is likely that patient age, arrival mode, and vital signs all latently encode patient frailty, which is known to be related to a patients reattendance risk^27^. Despite this, our predictive algorithm of choice, extreme gradient boosted decision trees, is relatively robust against correlations between variables and high variable redundancy is unlikely to be significantly detrimental to model performance.

In our exploratory analysis, we found that the hour of day the attendance began correlates to the reattendance rate, with higher reattendance rates observed during the night (Supplementary Figure 1). By inspection of the observed SHAP values for the hour of day (Figure 3 b), we observe that our model has learned a similar trend, associating attendance registration during the night with an increased (between zero and one percent increase) reattendance risk. This trend could have several different origins. Firstly, we have found (not shown) that the hour of day displays correlation with the reason for attendance, with either complaints associated with a higher (lower) risk of 72-hour reattendance more (less) likely to present during the night. Secondly, it is plausible that staff fatigue and lower staffing levels may contribute to the increased reattendance rate for attendances occurring during the night, although we have no way of testing this hypothesis in our dataset.

Our analysis (e.g., Figure 2 and model e in Table 2) shows that certain complaints are associated with a higher risk of 72-hour reattendance. For example, attendances whose chief complaint at registration is Abdominal pain had a mean 72-hour reattendance rate of 6.8 % (95 % CI: 6.0-7.7 %), compared to the mean reattendance rate of 4.8 % (95 % CI: 4.6-5.0 %) in the training set. Coding compliance was not evaluated in our dataset, which may effect this observation. For example, the most common chief complaint at registration was Unwell Adult (with 20.5 % of attendances listing this as the chief complaint in the training set) which is utilized when either the chief complaint is not clear at registration, when the patient presents with multiple complaints or as a result of inappropriate coding.

In addition to identifying complaints associated with a heightened short-term reattendance risk, our model also makes use of ICD10 coded conditions extracted from a patients electronic health record. These variables allow the model to identify medical conditions, comorbidities, and risks which are associated with increased reattendance risk and enables models to achieve moderate predictive performance (Table 2). Excluding the medical condition indicators, the most important feature is the 30-day visit count which, in part, reflects the disproportionate use of EDs by frequent users^28^. In the visualisation of the attendances in the patient hold-out test set in the two-dimensional explanation space (Figure 4), the most frequent attenders (30 day visit count of two or more) are clearly segregated (cluster 7 in Figure 4 b and Table 3). Visual inspection of the properties of each cluster in Figure 4 could be used to help guide the design of interventional strategies. For example, the reattendance risk of patients within cluster 7 (i.e., frequent attenders) could be reduced by increased provision of community support. Conversely, such support is unlikely to be the most appropriate intervention strategy for patients suffering from an acute injury (such as a severe burn) associated with increased reattendance risk.

From a clinical perspective it is interesting to investigate the subset of reattendances which are also readmissions (i.e., reattendances to the emergency department which result in subsequent admission to an inpatient ward). In these cases, there is increased risk that there was missed critical illness or injury at the initial attendance and these readmissions are important to evaluate for clinical assurance purposes. Overall, 37.1 % of reattendances end in readmission, resulting in a 72-hour readmission rate of 2.0 %. Evaluating our models predictions with a target equal to whether the patient readmitted in 72 hours, we find it has an AUROC of 0.804 (95 % CI: 0.774-0.832) and an average precision of 0.044 (95 % CI: 0.030-0.063) on the hold-out test set. The high AUROC means the model displays high discernibility between attendances which result in readmission and those that do not. The low average precision reflects that readmissions only make up a minority fraction of reattendances and the false positive rate increases as a result of the large class imbalance. Overall, these results demonstrate our model can identify the subset of reattendances which are also readmissions with a similar predictive performance as reattendances which do not result in admission, an important result since these two different outcomes will require different interventional strategies to reduce the risk of reattendance/readmission.

A limitation of our study, shared with other investigations of machine-learning use in EDs^29^, is that its primary data source is structured past medical histories. A medical history is not available for all patients and this could lead to our model discriminating against these patients. An example of this bias can be observed for cluster two (Table 3), where the two most important variables for determining a patients reattendance rate is the absence of any visit to the emergency department in the last 30 days and the absence of any recorded medical conditions. Unless implemented with consideration, using such a model could have adverse impact on patient care, because the model may not have sufficient descriptive information about a patient to make a reliable prediction of their reattendance risk. Patients who have several long-term health conditions, but have no past medical history with the hospital are particularly likely to be disadvantaged by this. We partially mitigate this bias by providing our model with visit-level information. In the future this bias could be reduced further by linking to community datasets (e.g., GP records) to obtain a view of a patients medical history, by providing confidence intervals alongside risk scores, and by educating clinicians on the limitations of the machine-learning model. In a deployment scenario, additional improvements could be achieved by using the model as an alert tool. In this setting, model predictions would only be presented to clinical staff for a small subset of patients; those the model predicts to be at particularly high risk of reattendance and for whom a decision to discharge has been made. Otherwise, the model would be invisible to clinical staff who would be free to carry out standard clinical practice in cases where an alarm is not raised.

Since our model only uses information available to clinicians at the time of the emergency department attendances it has a relatively low barrier to implementation. Despite this, it will be essential to perform prospective, randomized clinical trials of any implementation, investigating the efficacy of the model and the associated interventions. Ultimately, deployment of a machine-learning model could eventually invalidate the model by changing the behaviours and descriptors of reattendances by altering the clinical decisions made. In the short term, a relatively low-risk implementation of a model trained to identify patients at risk of reattendance would be in the implementation of a low-recall and high-precision alert system (for example, the configuration presented in Figure 2 c). This would only raise alarms for the cases the model believes are at the highest risk of reattendance and highlight the need for additional clinical review. On average, using the configuration displayed in Figure 2 c, this would have raised an alarm for only 1.7 % of attendances in which a decision to discharge was made (approximately 2 times per day) and would expect to be approximately 44 % of the time mitigating the risk of alarm fatigue. A model deployed in this manner would be of limited impact (because of its low recall), but the configuration could be re-evaluated if model performance improves. Performance could be improved by the inclusion of free-text notes recorded by clinical staff, which previous studies have shown can be predictive of patient outcomes across the broader hospital network^14,15^.

It is important to discuss the context in which our model could be prospectively deployed. Our model was trained and retrospectively evaluated using data available to clinicians during standard clinical practice at the emergency department in Southampton. This is an advantage if the model was to be used at this location because the characteristics of future attendances will likely reflect the attendances the model was trained on. Conversely, this means that the model will not necessarily generalize to other EDs without first training on their local data. Poor model generalization will be particularly prominent in EDs with a catchment zone with different demographics to Southampton and, therefore, different disease and illness prevalences. Because our model contains variables either in the standard UK emergency care dataset or regularly available to EDs nationally, it would be possible to evaluate this model directly in other EDs. External validation of our model is essential before prospective deployment beyond the department at which the training data was sourced.

## Conclusion

In conclusion, we have constructed and retrospectively evaluated an extreme gradient boosted decision tree model capable of predicting the 72-hour reattendance risk for a patient at the point of discharge from an emergency department. The highest performing model achieved an AUROC of 0.747 (95% CI : 0.722-0.773) and an average precision of 0.233 (95% CI : 0.194-0.277) on a hold-out test set. We investigated the variables most indicative of risk and showed these were patient level factors (medical history) rather than visit level variables such as recorded vital signs. We calculated SHAP values to explain our predictions (Figure 3) and used these to investigate the trends our model learned. We suggested an implementation of the algorithm in a low-recall, high-precision configuration such that alarms are only raised for patients predicted to be at a (clinically defined) heightened risk of reattendance. External validation and prospective clinical trials of our models is essential, with considerable consideration given to the planned interventions and the impact this would have on clinical decisions.

## Data Availability

The data that support the findings of this study are available from UHS, but restrictions apply to the availability of these data, which were used under license for the current study, and so are not publicly available. Data are however available from the authors upon reasonable request and with permission of UHS.

## Acknowledgements

This work was supported by The Alan Turing Institute under the EPSRC grant EP/N510129/1. We acknowledge support from the NIHR Wessex ARC.

## Author contributions statement

FPC and DKB performed the data analysis and modelling. ZDZ discussed and commented on the analysis with FPC and DKB. NW and FPC obtained governance and ethical approval. MA and FB created the data extract. FPC, MJB and NW managed the study at the UoS. MK managed the study at UHS. FPC and MK designed the study with assistance from TWVD. MK and TWVD provided clinical guidance and insight. FPC wrote the first draft of the manuscript with assistance from MK and TWVD. All authors frequently discussed the work and commented and contributed to future drafts of the manuscript.

## Additional information

### Data availability

Due to patient privacy concerns the dataset used in this study is not publicly available. However, it will be made available upon reasonable request

## References

1. Berchet, C., Emergency care services: trends, drivers and interventions to manage the demand. (2015)

2. Baier, N., et al. Emergency and urgent care systems in Australia, Denmark, England, France, Germany and the Nether-landsAnalyzing organization, payment and reforms. Health Policy 123, 1–10 (2019)

3. Bernstein, S. L., et al. The effect of emergency department crowding on clinically oriented outcomes. Academic Emergency Medicine 16, 1–10 (2009)

4. Guttmann, A., et al. Association between waiting times and short term mortality and hospital admission after departure from emergency department: population based cohort study from Ontario, Canada. BMJ 342, d2983 (2011)

5. Besga, A., et al. Risk factors for emergency department short time readmission in stratified population. BioMed research international (2015)

6. Deschodt, M., et al. Characteristics of older adults admitted to the emergency department (ED) and their risk factors for ED reattendance based on comprehensive geriatric assessment: a prospective cohort study. BMC geriatrics 15, 1 (2015)

7. Martin-Gill, C., Reiser, R.C. Risk factors for 72-hour admission to the ED. The American journal of emergency medicine 22 6, 448–453 (2004)

8. Arendts, G., Fitzhardinge, S., Pronk, K., Hutton, M., Nagree, Y. and Donaldson, M. Derivation of a nomogram to estimate probability of revisit in at-risk older adults discharged from the emergency department. Internal and emergency medicine 8 3, 249–254 (2013)

9. Kun-Hsing, Y., Beam A. L. and Kohane, I. S. Artificial intelligence in healthcare. Nature biomedical engineering 2, 10, 719–731 (2018)

10. Moncada-Torres, A., van Maaren, M.C., Hendriks, M.P., Siesling, S. and Geleijnse, G. Explainable machine learning can outperform Cox regression predictions and provide insights in breast cancer survival. Scientific Reports 11, 1, 1–13 (2021)

11. Li, Y., et al. An Interpretable Machine Learning Survival Model for Predicting Long-term Kidney Outcomes in IgA Nephropathy. In AMIA Annual Symposium Proceedings 737 (2020)

12. Hao. S., et al. Risk prediction of emergency department revisit 30 days post discharge: a prospective study. PLoS one 9 11, e112944 (2014)

13. Hong, W.S., Haimovich, A.D. and Taylor, R.A. Predicting 72-hour and 9-day return to the emergency department using machine learning. JAMIA open 2 3, 346–352 (2019)

14. Huang, K., Altosaar J. and Ranganath R. Clinicalbert: Modeling clinical notes and predicting hospital readmission. arXiv, 1904.05342 (2019)

15. Sterling, N. W., Patzer, R. E., Di, M., and Schrager, J. D. Prediction of emergency department patient disposition based on natural language processing of triage notes. International journal of medical informatics, 129, 184–188 (2019)

16. Micci-Barreca, D. A preprocessing scheme for high-cardinality categorical attributes in classification and prediction problems. ACM SIGKDD Explorations Newsletter, 3 1, 27–32 (2001)

17. Bergstra, J.S., Bardenet, R., Bengio, Y. and Kégl, B. Algorithms for hyper-parameter optimization. In Advances in neural information processing systems, 2546–2554 (2011)

18. Bergstra, J., Yamins, D., and Cox, D. D. Making a Science of Model Search: Hyperparameter Optimization in Hundreds of Dimensions for Vision Architectures. In International Conference on Machine Learning, 115–123 (2013)

19. Chen, T. and Guestrin, C. XGBoost: A scalable tree boosting system. In Proceedings of the 22nd ACM sigkdd international conference on knowledge discovery and data mining, 785–794 (2016)

20. Lundberg, S. M., and Su-In L. A unified approach to interpreting model predictions. In Advances in neural information processing systems, 4765–4774 (2017)

21. Lundberg, S. M., et al. From local explanations to global understanding with explainable AI for trees. Nature machine intelligence 2 1, 2522–5839 (2020)

22. Lundberg, S. M., et al. Explainable machine-learning predictions for the prevention of hypoxaemia during surgery. Nature biomedical engineering 2 10, 749–760 (2018)

23. McInnes, L., Healy, J. UMAP: Uniform Manifold Approximation and Projection for Dimension Reduction, arXiv 2 1802.03426 (2018)

24. Becht, E., et al. Dimensionality reduction for visualizing single-cell data using UMAP. Nature biotechnology 37 1,38–44 (2019)

25. Diaz-Papkovich, A., Anderson-Trocmé, L., Gravel, S. UMAP reveals cryptic population structure and phenotype heterogeneity in large genomic cohorts. PLoS genetics 15 11, e1008432 (2019)

26. Ester, M., Kriegel, H.P., Sander, J. and Xu, X. A density-based algorithm for discovering clusters in large spatial databases with noise. In Kdd 96 34, 226–231 (1996)

27. Kahlon, S., et al. Association between frailty and 30-day outcomes after discharge from hospital. Cmaj 187 11, 799–804 (2015)

28. LaCalle, E. and Rabin, E. Frequent users of emergency departments: the myths, the data, and the policy implications. Annals of emergency medicine 56 1, 42–48 (2010)

29. Joseph, J.W., et al. Deep-Learning Approaches to Identify Critically Ill Patients at Emergency Department Triage Using Limited Information. JACEP Open 1, 773–781 (2020)

