## Supplementary Information for "Using explainable machine learning to identify patients at risk of reattendance at discharge from emergency departments"

### Predictive variables

| Variable | Data type | Encoding | Description |
|---|---|---|---|
| Age | Integer | - | Age of patient at attendance. Estimated from year of attendance minus year of birth. |
| Complaint (attendance) | Categorical | Target | Chief complaint (coded) for attendance recorded at registration. |
| Complaint (triage) | Categorical | Target | Chief complaint (coded) for attendance recorded at triage. |
| Diagnosis | Categorical | Target | Primary diagnosis at point of discharge. |
| Discriminator | Categorical | Target | Discriminator (e.g., 'Viral Wheeze') recorded at triage. |
| Manchester Triage System score | Integer | - | Result of the Manchester Triage System (1-5). |
| Pain score | Integer | - | Pain score (scale of 0-10) recorded at triage. |
| Arrival mode | Categorical | Target | Mode of arrival (e.g., Emergency road ambulance). |
| Condition count | Integer | - | Count of conditions/ risk mentionedin all patients' inpatient medical discharge summaries prior to current attendance. |
| Condition indicator | Categorical | One-hot | Binary indicator of whether patient has history of a given condition/risk (e.g., hypertension, current smoker, type 2 diabetes). |
| 30-day visit count | Integer | - | Number of emergency department attendances the patient has made in the last 30 days. |
| Temperature (vital signs) | Continuous | - | Temperature of patient, measured at triage. |
| Systolic blood pressure (vital signs) | Continuous | - | Systolic blood pressure, measured at triage. |
| Respiration rate (vital signs) | Integer | - | Respiration rate, visually measured at triage. |
| Pulse rate (vital signs) | Integer | - | Pulse rate, measured at triage. |
| Blood oxygen saturation (vital signs) | Continuous | - | Blood oxygen saturation, measured at triage. |
| Hour of day (temporal) | Integer | - | Hour of day (0-23) patient registered for attendance. |
| Day of week (temporal) | Categorical | Nominal | Day of week (0-6) patient registered for attendance. |

**Supplementary Table 1: Variables used in our modelling and high-level descriptions of them.** The encoding scheme used for categorical variables is displayed in the third column.

| Variable | Data type | Encoding | Mean | Mode | Missing fraction |
|---|---|---|---|---|---|
| Age | Integer | - | 46.5 years | 21 years | 0.0 |
| Complaint (attendance) | Categorical | Target | - | - | 0.0 |
| Complaint (triage) | Categorical | Target | - | Limb problem | 0.0 |
| Diagnosis | Categorical | Target | - | No abnormality detected | 0.0 |
| Discriminator | Categorical | Target | - | Recent problem | 0.0. |
| Manchester Triage System score | Integer | - | 3.3 | 3 | 0.0 |
| Pain score | Integer | - | 2.6 | 0 | 0.0 |
| Arrival mode | Categorical | Target | - | Patient walk-in | 0.0 |
| Condition count | Integer | - | 1.3 | 0 | 0.0 |
| Condition indicators | Categorical | One-hot | - | - | - |
| 30-day visit count | Integer | - | 0.3 | 0 | 0.0 |
| Temperature (vital signs) | Continuous | - | 36.7 | - | 66% |
| Systolic blood pressure (vital signs) | Continuous | - | 139.4 | - | 66% |
| Respiration rate (vital signs) | Continuous | - | 18.3 | - | 66% |
| Pulse rate (vital signs) | Continuous | - | 82.8 | - | 66% |
| Blood oxygen saturation (vital signs) | Continuous | - | 97.2 | - | 66% |
| Hour of day (temporal) | Integer | - | 13.4 | 11 | 0.0 |
| Day of week (temporal) | Categorical | Nominal | - | Monday | 0.0 |

**Supplementary Table 2: Variables used in our modelling and quantitative descriptors of them.** 66 % of attendances did not have associated vital signs recorded. This could of been because it was not deemed necessary by clinical staff, their condition was particularly severe, or because they were not recorded.

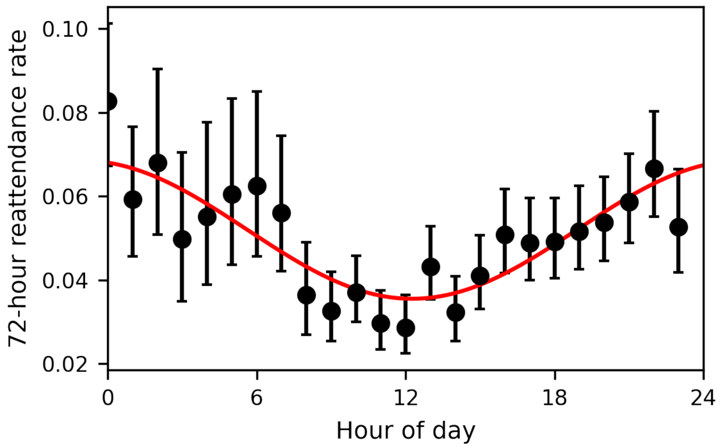


**Supplementary Figure 1: Observed 72-hour reattendance rate as a function of the hour of day the patient registered for the attendances.** Black markers denote observed reattendance rate, error bars are 95 % confidence intervals calculated using the Wilson score interval. The red solid line is a cosine fit to the data ($f(x) = A\cos\left(B(x-C)\right)+D$) to demonstrate the periodic nature of the observed reattendance rate.

### Modelling

| Parameters | Value | Description |
|---|---|---|
| colsample_bytree | 0.786 | Fraction of variables to subsample for each tree. |
| gamma | 4.618 | Regularisation parameter - the minimum loss reduction required to make a further partition on a leaf node. |
| learning_rate | 0.038 | Step size shrinkage used in weight update. Smaller value make the boosting more conservative. |
| max_delta_step | 2.381 | Maximum delta step of each trees weight. Used to make the boosting process more conservative. |
| max_depth | 5 | Maximum depth of a tree. |
| n_estimators | 96 | Number of boosting iterations. |
| subsample | 0.613 | Subsample ratio of the training instances, performed at each boosting iteration. |

**Supplementary Table 3: Hyperparameters for our final reattendance model using all variables available to it at the time of discharge.**

### SHAP values and local explainability

By using the TreeExplainer algorithm we calculated the SHAP values for our XGBoost model. These values provide predictions at an instance level, meaning that for a given attendance we can present a predicted reattendance risk score and break down this prediction at the variable level, assigning each variable a scalar value (i.e., a SHAP value) which reflects the impact (in terms of both magnitude and direction) that variable had on model predicting the given risk score. Providing a breakdown of a a patients reattendance risk at a variable level, could allow the design of bespoke interventions based on both the predicted risk and the reason for this risk.

In Supplementary Figure 2 we show the local explanation for a entirely synthetic patient, where each variable was set to the mode value of all patients in the dataset, with the exception of medical history where the synthetic patient was set to have a history of type 1 diabetes and a history of smoking and the patient was set to have an age of 40 years old. Supplementary Figure 2 displays the five most important variables for this synthetic patient (as determined by the magnitude of the SHAP values), each bar displays how a given feature changes this patients risk relative to the baseline risk. For example, the patient has had no visits to the emergency department in the past 30-days and the model associates this with a reduced risk of reattendance (solid blue bar) relative to the baseline reattendance risk. However, the patient has a history of smoking and the model associates this with an increased reattendance risk.

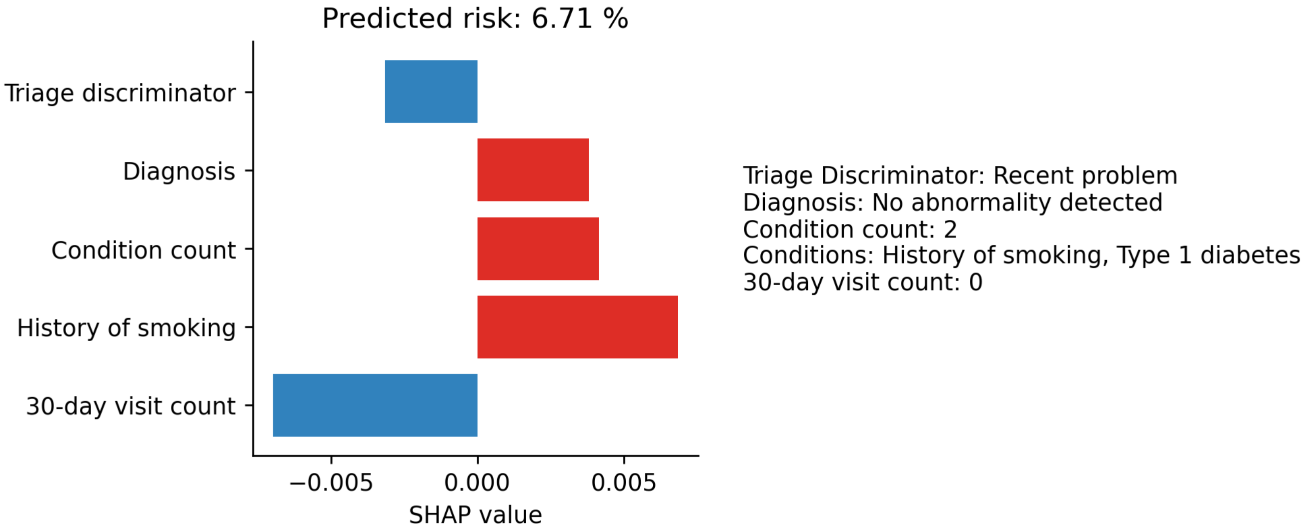


**Supplementary Figure 2: An explanation of a synthetic patient's reattendance risk.** The graphic displays the SHAP values for the five most important features in determining this patients reattendance risk. The model associates two variables (Triage discriminator and 30-day visit count - blue bars) with a reduction in the patients reattendance risk relative to the baseline risk and associates three variables (Diagnosis, condition count, history of smoking - red bars) with an increased relatives risk of reattendance.
